# Functional Profiling of Six *KCNJ2* Variants Reveals Distinct Dominant-Negative and Haploinsufficiency Mechanisms in Andersen-Tawil Syndrome

**DOI:** 10.64898/2026.11.27.26367880

**Authors:** Takafumi Iguchi, Seiko Ohno, Futoshi Toyoda, Takanori Aizawa, Takeru Makiyama, Minoru Horie, Ken Terata, Yoko Yoshida, Akira Mizukami, Naokata Sumitomo, Kisei Endo, Wataru Shimizu, Takeshi Aiba, Koichi Kato

**Author notes:** Corresponding Author: Koichi Kato, MD, PhD, Department of Cardiovascular Medicine, Shiga University of Medical Science Seta, Tsukinowa-cho, Otsu, Shiga 520-2192, Japan.

## Abstract

**BACKGROUND:** Andersen-Tawil syndrome (ATS) is caused by pathogenic *KCNJ2* variants and is characterized by ventricular arrhythmias, periodic paralysis, and dysmorphic features, occasionally leading to sudden death. These heterozygous variants encode the Kir2.1 channel, which contributes to the maintenance of the cardiomyocyte resting membrane potential and repolarization. Although many variants cause loss of function (LOF) through dominant-negative (DN) effects or haploinsufficiency, the functional consequences of some variants remain unknown. In this study, we aimed to elucidate the pathogenicity and molecular mechanisms of uncharacterized *KCNJ2* variants.

**METHODS:** From the NCVC and SUMS cohorts, six uncharacterized variants in *KCNJ2* (p.I72T, p.76insT, p.103_104del, p.F135S, p.N190I, and p.Q310R) were evaluated. In HEK293 cells, macroscopic currents and DN effects were assessed by whole-cell patch-clamp. Channel localization and trafficking were assessed by confocal microscopy. Specifically, for the p.Q310R variant, co-immunoprecipitation was performed to assess co-assembly with the wild-type (WT) subunits.

**RESULTS:** The six variants exhibited distinct patterns of channel dysfunction. The p.103_104del and p.F135S variants showed severe trafficking defects with decreased membrane accumulation and were associated with strong DN effects. The p.76insT and p.N190I variants showed preserved membrane trafficking despite strong DN effects. The p.I72T variant retained partial channel function and exhibited normal trafficking. Notably, despite normal trafficking and preserved co-assembly with WT subunits to form multimers, the p.Q310R variant showed a functional pattern consistent with haploinsufficiency, without an apparent DN effect. Clinically, carriers of variants lacking a DN effect (p.Q310R) or retaining partial independent function (p.I72T) appeared to have fewer classical features of the syndrome, such as absent dysmorphism.

**CONCLUSIONS:** *KCNJ2* variants cause LOF through diverse mechanisms, including impaired trafficking and functional defects despite preserved trafficking. Specifically, the p.Q310R variant shows loss of channel function without a detectable DN effect despite preserved co-assembly with WT subunits. Differences in residual channel function may contribute to phenotypic variability.

**Clinical Perspective:** *What is known?:*

- Andersen-Tawil syndrome is primarily caused by loss-of-function variants in *KCNJ2*, which encodes the Kir2.1 potassium channel.
- These pathogenic variants lead to a loss of channel function in the form of a dominant-negative effect or haploinsufficiency through mechanisms such as trafficking defects or pore dysfunction.

*What the study adds:*

- Comprehensive profiling of six uncharacterized *KCNJ2* variants revealed diverse functional consequences, including dominant-negative effects caused by trafficking defects, pore dysfunction despite normal trafficking, rare variants presenting with typical haploinsufficiency despite normal trafficking (p.Q310R), or retaining partial channel function even as mutant homotetramers (p.I72T).
- Patients carrying variants with residual independent function (p.I72T) or lacking a dominant-negative effect (p.Q310R) tended to present with incomplete classical features of the syndrome, suggesting that residual channel function may influence clinical phenotypic variability.

## INTRODUCTION

Andersen-Tawil syndrome (ATS) is a rare autosomal dominant disorder characterized by ventricular arrhythmias, periodic paralysis, and dysmorphic features^1)^. Most cases arise from heterozygous variants in *KCNJ2* (ATS type 1), which encodes the inward rectifier potassium channel Kir2.1, a critical regulator of cardiomyocyte resting membrane potential and repolarization^2)3)^. Over 70 pathogenic or likely pathogenic *KCNJ2* variants have been reported, with patch-clamp analyses reported for at least 50 of the variants (ClinVar database, accessed Oct 2026)^3)4)5)^. These studies demonstrate that ATS-causing mutant channels predominantly exhibit a loss-of-function (LOF) phenotype. This LOF typically manifests as either a dominant-negative (DN) effect or haploinsufficiency, driven by diverse cellular mechanisms including intracellular trafficking defects.

The Japanese long-QT syndrome (LQTS) registry is a nationwide registry including almost 4,000 patients with congenital LQTS and LQTS-associated syndromes, such as ATS^6)^. In the registry, we identified a total of 113 patients with ATS carrying 36 distinct *KCNJ2* variants. Although the functional properties of most of these variants have been well characterized in previous studies^7–15)^, several variants have not yet been functionally validated. In this study, to characterize their functional effects and underlying mechanisms, we used a mammalian cell expression system and whole-cell patch-clamp recordings to evaluate current densities and DN effects, alongside confocal microscopy to identify trafficking abnormalities.

## METHODS

### *KCNJ2* Uncharacterized Variants in ATS

From the NCVC and SUMS inherited arrhythmia cohorts, a total of twelve ATS patients with six *KCNJ2* variants (c.215T>C [p.I72T], c.222_224dupCAC [p.76insT], c.309_314delTTGGTT [p.103_104del], c.404T>C [p.F135S], c.569A>T [p.N190I], and c.929A>G [p.Q310R]) which were not functionally characterized by the molecular and physiological studies were enrolled.

### Clinical Data Collection and Ethical Approval

The data that support the findings of this study are available from the corresponding author upon reasonable request. This study was approved by the institutional review boards of Shiga University of Medical Science (Approval Nos. G2011-128) and the National Cerebral and Cardiovascular Center Ethics Committee (Approval Nos. R22006-10 and M29-167-11). The investigation was conducted in accordance with the principles outlined in the Declaration of Helsinki. All patients or their legal guardians provided written informed consent prior to the clinical and genetic investigations. Detailed clinical characteristics and electrocardiographic data of the variant carriers were retrospectively collected through questionnaire surveys completed by their treating physicians.

### Plasmid Construction and Mutagenesis

For electrophysiological analyses, a previously described plasmid encoding full-length wild-type (WT) *KCNJ2* in the pCMS-EGFP expression vector (Clontech, Palo Alto, CA, USA) was used as a template^15)^. Six *KCNJ2* variants (p.I72T, p.76insT, p.103_104del, p.F135S, p.N190I, and p.Q310R) were introduced into the WT construct utilizing the QuikChange II XL site-directed mutagenesis kit (Stratagene, La Jolla, CA, USA). The presence of each intended variant was confirmed using direct Sanger sequencing.

For immunocytochemical evaluations, the WT and *KCNJ2* variant constructs generated for patch-clamp experiments were transferred into the pcDNA3.1(+) vector (Thermo Scientific, Waltham, MA, USA) using the In-Fusion HD Cloning Kit (Takara Bio, Shiga, Japan). To assess cell-surface expression of the channels, a hemagglutinin (HA) epitope tag (YPYDVPDYA) was engineered into the extracellular loop positioned between the first and second transmembrane segments (TM1 and TM2) of the Kir2.1 protein, specifically inserting it between the Ala-115 and Ser-116 residues^15)^.

### Cell Culture and Transient Transfection

Human embryonic kidney 293 (HEK293) cells were utilized^10)^. The cells were maintained in Dulbecco’s Modified Eagle Medium (DMEM) containing 4.5 g/L glucose, supplemented with 5% fetal bovine serum (FBS) and a 1% penicillin/streptomycin mixture (Nacalai Tesque Inc., Kyoto, Japan). The cells were cultured in a humidified incubator at 37°C with a 5% CO_2_ atmosphere. Plasmids were transiently transfected into the HEK293 cells using the jetPEI reagent (Polyplus-transfection, Illkirch, France) according to the manufacturer’s instructions. Cells seeded in 35-mm dishes received 0.5 μg of plasmid DNA for whole-cell patch-clamp recordings and 1.0 μg for immunocytochemistry assays. For co-immunoprecipitation (Co-IP) assays, cells were cultured in 60-mm dishes and co-transfected with 3.0 μg of each plasmid DNA.

### Electrophysiological Analysis

Macroscopic Kir2.1 currents were recorded using the conventional whole-cell patch-clamp technique 36 to 60 hours post-transfection. Only cells with robust green fluorescent protein (GFP) signals were selected for recording. All electrophysiological measurements were performed at 37°C using an EPC-8 patch-clamp amplifier (HEKA Elektronik, Lambrecht, Germany).

To elicit Kir2.1 currents, the membrane potential was held at -80 mV and subsequently stepped from -140 mV to +20 mV in 10-mV increments for a duration of 500 ms. The borosilicate glass pipettes were filled with an internal solution consisting of (in mM): 70 K-aspartate, 70 KOH, 40 KCl, 10 KH_2_PO_4_, 1 MgSO_4_, 3 K_2_-ATP, 5 EGTA, 0.1 Li_2_-GTP, and 5 HEPES (pH adjusted to 7.2 with KOH). The extracellular bath solution contained (in mM): 140 NaCl, 5.4 KCl, 0.5 MgCl_2_, 1.8 CaCl_2_, 0.33 NaH_2_PO_4_, 5.5 glucose, and 5 HEPES (pH adjusted to 7.4 with NaOH)^15)^.

### Immunocytochemistry

To evaluate the subcellular localization and membrane trafficking of the channels, immunofluorescence staining was performed. At 48 hours post-transfection, the cultured HEK293 cells were washed twice with phosphate-buffered saline (PBS). Subsequently, the samples were treated with a rabbit anti-HA primary antibody (1:200 dilution; Cell Signaling Technology, Danvers, MA, USA) and incubated overnight at 4°C. After two further washes with PBS, the cells were incubated with an Alexa Fluor 488-conjugated goat anti-rabbit secondary antibody (diluted 1:1000 in PBS supplemented with 2% FBS) for 120 minutes at room temperature. Following a final PBS wash, the cells were mounted using Mounting Medium with DAPI (Sigma-Aldrich, St. Louis, MO, USA) to counterstain the nuclei. The cellular localization of the targeted channels was then examined using a TCS SP8 X confocal laser scanning microscope (Leica, Wetzlar, Hessen, Germany).

### Co-immunoprecipitation and Western Blotting

To examine channel subunit interactions following the functional and trafficking analyses, a Co-IP assay was performed to determine whether the Q310R variant can co-assemble with WT channel subunits. HEK293 cells seeded in 60-mm dishes were co-transfected with FLAG-tagged WT combined with either non-tagged WT, HA-tagged WT, or HA-tagged Q310R. At 48 hours post-transfection, cell lysates were prepared and incubated with an anti-HA antibody (1:200 dilution) overnight on a rotator at 4°C. The protein complexes were subsequently isolated using Protein G Dynabeads.

The eluted samples were mixed with a sample loading buffer and resolved by gel electrophoresis at 100 V for 150 minutes. Separated proteins were then electrotransferred onto PVDF membranes utilizing the Trans-Blot Turbo blotting system (BIO-RAD, Berkeley, CA, USA) at 1.0 V (25 mA) for 30 minutes. Antibody incubations were performed using the iBind Flex Western System (Invitrogen, Carlsbad, CA, USA). The membranes were probed with a primary anti-FLAG antibody (F3165; Sigma, USA) at a 1:350 dilution and a rabbit anti-GAPDH antibody at a 1:1000 dilution. For detection, an anti-mouse secondary antibody (1:1000 dilution) and the Clean-Blot IP Detection Kit (1:1000 dilution) were employed. Finally, protein signals were visualized using a chemiluminescence reagent (Chemi-Lumi One; Nacalai Tesque, Kyoto, Japan) and digitally captured with an ImageQuant LAS-4000 imager (Cytiva, Marlborough, MA, USA).

### Image Analysis and Statistical Analysis

To quantify the membrane accumulation of the channels, confocal images were analyzed using Fiji software^16)^. The plasma membrane region was strictly defined as a region extending 1 μm inward from the cell surface^17)^. Membrane-trafficking efficiency was then evaluated by calculating the ratio of fluorescence intensity at the defined membrane region to that within the cytoplasm. Electrophysiological data are presented as mean ± standard error of the mean (SEM). All statistical analyses were conducted using R software (version 4.4.1). For the evaluation of patch-clamp electrophysiological data, Welch’s t-test was used. To assess differences in the membrane accumulation ratios across the groups, groups were compared using a one-way analysis of variance (ANOVA) followed by Dunnett’s multiple comparison test, with the WT serving as the control group. For the quantitative analysis of co-immunoprecipitation, the Kir2.1/GAPDH ratio of each group was normalized to that of the HA-WT group from the same independent experiment to control for inter-assay variability. Statistical significance of the normalized co-immunoprecipitation data was determined using a one-sample t-test against a hypothetical value of 1.0. Statistical significance thresholds were set at *p* < 0.05 and *p* < 0.01.

## RESULTS

### Clinical Characteristics

Twelve patients from the Japanese LQTS registry carrying one of the six uncharacterized *KCNJ2* variants were included in this study. The clinical characteristics and clinical phenotypes of these patients are summarized in **Table 1**. The I72T and the 103_104del variant were each identified in only one patient, whereas multiple carriers were identified for the other four variants.

**Table 1.** Clinical Characteristics of ATS Patients with Uncharacterized *KCNJ2* Variant.

| Case No. | Age range | Sex | Proband/Family | Nucleotide change | Amino acid change | Variant location | ECG | QTc (ms) | QUc (ms) | Dysmorphism | Paralysis | Syncope/LAE | Treatment |
|---|---|---|---|---|---|---|---|---|---|---|---|---|---|
| 1 | 30-34 | F | Proband | 215T>C | I72T | N | 1, 4 | 427 | 642 | - | - | -/- | BB |
| 2 | 5-9 | F | Proband | 222-224dupCAC | 76insT | N | 1, 2 | 384 | 727 | + | + | -/- | FL |
| 3 | 35-39 | F | Family | 222-224dupCAC | 76insT | N | - | 523 | 694 | - | - | -/- | - |
| 4 | 0-4 | M | Family | 222-224dupCAC | 76insT | N | 1 | 377 | 699 | + | + | -/- | - |
| 5 | 20-24 | F | Proband | 309_314delTTGGTT | 103_104del | Pore | 1 | 428 | 688 | + | - | -/- | BB, FL |
| 6 | 15-19 | F | Proband | 404T>C | F135S | Pore | 1, 3 | NA | NA | + | + | -/- | BB, FL, IV |
| 7 | 15-19 | M | Family | 404T>C | F135S | Pore | 1, 3 | 415 | 631 | + | - | -/- | FL, IV |
| 8 | 10-14 | F | Proband | 569A>T | N190I | C | 1, 2 | 445 | 615 | + | - | -/- | BB, FL |
| 9 | 40-44 | F | Family | 569A>T | N190I | C | 1, 3 | 415 | 631 | + | - | -/- | - |
| 10 | 10-14 | F | Proband | 929A>G | Q310R | C | 1, 3 | 547 | 732 | - | + | -/- | - |
| 11 | 45-49 | F | Family | 929A>G | Q310R | C | 1 | 470 | 695 | - | - | -/- | - |
| 12 | 5-9 | M | Family | 929A>G | Q310R | C | - | 398 | 623 | - | - | -/- | - |

Age range; age range at diagnosis, LAE=lethal arrhythmic event, NA=not available, N=N-terminal, C=C-terminal, BB=β-blocker, FL=flecainide, Ⅳ=verapamil, ECG: 1=PVC, 2=bidirectional VT, 3=monomorphic VT, 4=polymorphic VT

Clinical presentations varied among the variant carriers. The patient with the I72T variant presented with ventricular arrhythmias alone (premature ventricular contractions and polymorphic ventricular tachycardia) without any extracardiac features. Among the three patients harboring the Q310R variant, the clinical presentation was heterogeneous: one case exhibited both periodic paralysis and ventricular arrhythmias, another presented with isolated ventricular arrhythmias, and the third case displayed only an isolated abnormal U wave without documented arrhythmias or paralysis. Importantly, characteristic U waves were observed on the electrocardiograms of all patients. Notably, despite the presence of repolarization abnormalities and ventricular arrhythmias, none of the patients in this cohort experienced syncope or lethal arrhythmic events.

### Electrophysiological Functional Analysis

The topological locations of the six uncharacterized *KCNJ2* variants within the Kir2.1 channel subunit are illustrated in **Figure 1A**. To assess the functional properties of all variants, WT or each variant plasmid was expressed alone (0.5 µg) in HEK293 cells following the methods described above. Representative macroscopic current traces are shown in **Figure 1B-a**. Consistent with typical Kir2.1 channel properties, WT exhibited strong inward rectification, as depicted in the current-voltage (I-V) relationship in **Figure 1C**. The I72T variant exhibited partial function, generating 42% of the inward current at -130 mV and 14% of the outward current at -60 mV relative to the WT. In contrast, all other variants yielded no measurable currents, indicating a complete loss of function. Next, to mimic the heterozygous state of the alleles in patients, WT and each *KCNJ2* variant plasmids were co-transfected at an equimolar ratio (0.25 µg each). For the WT-alone control (0.25 µg), the total amount of plasmid was adjusted to 0.5 µg using the pEGFP-C3 vector. The resulting current traces for the co-transfections are shown in **Figure 1B-b**.

**Figure 1.** Electrophysiological properties of wild-type (WT) and mutant Kir2.1 channels expressed in HEK293 cells. (A) Schematic topology of the Kir2.1 channel subunit indicating the locations of the six KCNJ2 variants analyzed in this study. (B) Representative whole-cell macroscopic current traces of WT and variants expressed alone (0.5 μg) (a) or co-expressed with WT (0.25 μg + 0.25 μg) to mimic the heterozygous state (b). Currents were elicited by 500-ms voltage steps from a holding potential of -80 mV to test potentials ranging from -140 mV to +20 mV in 10-mV increments, as illustrated in the inset. (C) Current-voltage (I-V) relationships of WT and variants. (D) Bar graphs showing peak inward current densities at -130 mV and peak outward current densities at -60 mV for each group. Error bars represent the standard error of the mean. * indicates statistical significance compared with the WT control evaluated by Welch’s t-test. † indicates a current density of less than 75% relative to the half-WT (control).

The peak current densities at -130 mV and -60 mV for each co-transfected group are summarized in Table 2 and presented as bar graphs in **Figure 1D**. When co-expressed with WT, the 76insT, 103_104del, F135S, and N190I variants yielded 49%/53% (at -130 mV / at -60 mV), 55%/60%, 47%/61%, and 52%/70% of the current densities compared to the WT (0.25 µg) control, respectively. Defining a dominant-negative effect as a reduction in current density to less than 75% of the WT (0.25 µg) control, the 76insT, 103_104del, F135S, and N190I variants exerted a clear dominant-negative suppression effect on both inward and outward Kir2.1 currents^18)^. Conversely, I72T demonstrated partial function without suppressing the WT, and Q310R exhibited a haploinsufficiency effect.

**Table 2.** Peak current densities of co-expressed mutant channels.

| Co-expressed | n | Inward Current at -130 | % of WT | Outward Current at -60 | % of WT |
|---|---|---|---|---|---|
| WT (250 ng) | 25 | -283.7 ± 55.1 | 100% | 16.2 ± 3.5 | 100% |
| I72T | 15 | -302.5 ± 24.3 | 107% | 20.9 ± 5.9 | 130% |
| 76insT | 25 | -138.8 ± 19.4<sup>†</sup> | 49% | 8.6 ± 2.8<sup>†</sup> | 53% |
| 103_104del | 14 | -155.2 ± 24.9<sup>†</sup> | 55% | 9.7 ± 1.3<sup>†</sup> | 60% |
| F135S | 23 | -134.5 ± 26.3<sup>†</sup> | 47% | 9.9 ± 1.8<sup>†</sup> | 61% |
| N190I | 15 | -148.0 ± 28.8<sup>†</sup> | 52% | 11.3 ± 3.1<sup>†</sup> | 70% |
| Q310R | 19 | -244.7 ± 34.1 | 86% | 15.0 ± 2.3 | 93% |

Data are shown as mean ± standard error. Current densities were measured at -130 mV for inward currents and -60 mV for outward currents. The relative current percentage (% of WT) was calculated against the wild-type (WT) 250 ng control group. † indicates a current density of less than 75% relative to the half-WT (control)

### Intracellular Trafficking Assessment: Immunocytochemistry

To evaluate the subcellular localization and membrane trafficking of the mutant Kir2.1 channels, we acquired images using a confocal laser scanning microscope targeting the HA-tagged channels. Representative images are shown in **Figure 2A**. The WT, I72T, 76insT, N190I, and Q310R channels demonstrated normal forward trafficking and distinct localization to the plasma membrane. In contrast, the 103_104del and F135S variants lacked clear peripheral fluorescence outlining the cell membrane, revealing significant intracellular trafficking errors. **Figure 2B** quantifies the membrane-to-cytoplasm accumulation ratio for each group. Consistent with the visual findings in **Figure 2A**, the two variants exhibiting trafficking defects (103_104del and F135S) showed statistically significant reductions in membrane accumulation compared to the WT control.

**Figure 2.** Subcellular localization and membrane trafficking of WT and mutant Kir2.1 channels. (A) Representative confocal immunofluorescence images of HEK293 cells transfected with HA-tagged WT or mutant Kir2.1 channels. Green fluorescence (HA-Ab) indicates Kir2.1 channels, and blue fluorescence (DAPI) indicates nuclei. Scale bar = 25 μm. (B) Quantitative analysis of membrane trafficking efficiency, calculated as the ratio of fluorescence intensity at the plasma membrane to that in the cytoplasm. Data are shown as box plots with individual data points (n = 20–21 cells per group). * *p* < 0.05 vs. WT control (one-way ANOVA followed by Dunnett’s multiple comparison test).

### Co-immunoprecipitation Assay for the Q310R Variant

Our initial assessments revealed that the Q310R variant traffics normally to the cell membrane but lacks a DN effect. To further elucidate its intracellular behavior and determine whether Q310R retains the capability to physically interact and form heteromeric tetrameric channels with WT Kir2.1, we performed Co-IP assays using FLAG-tagged WT and HA-tagged Q310R. Following precipitation with an anti-HA antibody, the samples were immunoblotted using an anti-FLAG antibody to detect chemiluminescence. A representative Co-IP result is shown in **Figure 3A**, confirming that non-HA-tagged WT channels were barely precipitated (negative control). Notably, HA-Q310R was able to co-precipitate FLAG-WT at a yield closely equivalent to that of HA-WT.

**Figure 3.** The Q310R variant maintains the ability to co-assemble with WT Kir2.1. (A) Representative co-immunoprecipitation (Co-IP) and total-protein Western blot images from HEK293 cells co-expressing FLAG-WT and either non-tagged WT (negative control), HA-WT, or HA-Q310R. Proteins were immunoprecipitated with an anti-HA antibody and detected with an anti-FLAG antibody. (B) Quantitative analysis of co-immunoprecipitated Kir2.1. The Kir2.1/GAPDH ratio of each group was normalized to that of the HA-WT group (set to 1.0, indicated by the dashed line) from the same independent experiment to control for inter-assay variability. Data are presented as box plots with individual data points (n = 5 independent experiments). *p* < 0.01 vs. HA-WT (one-sample t-test); ns, not significant.

**Figure 3B** summarizes the quantitative results from 5 independent experiments, presented as box plots. To control for inter-assay variability, the Kir2.1/GAPDH ratio of each group was normalized to that of the HA-WT group (set to 1.0) from the same independent experiment. There was no statistically significant difference between HA-Q310R and the hypothetical value of 1.0 (HA-WT). These findings indicate that the Q310R mutation does not impair the channel’s ability to physically co-assemble with WT Kir2.1 subunits.

## DISCUSSION

### Overview and Significance of the Study

In this study, we conducted electrophysiological and cell biological functional analyses of six *KCNJ2* variants (I72T, 76insT, 103_104del, F135S, N190I, and Q310R) identified from the Japanese LQTS registry, whose functional characteristics were previously unknown. The results revealed that these variants cause loss of function (LOF) through diverse molecular mechanisms, including intracellular trafficking defects, loss of pore function, and a unique haploinsufficiency. Since variants in *KCNJ2* were first identified as the cause of ATS^2)^, progress has been made in elucidating the pathophysiology at the molecular level; the findings of this study further deepen our understanding of the detailed mechanisms underlying the dysfunction of each variant.

### Dominant-Negative Effect Due to Trafficking Defects (103_104del, F135S)

The 103_104del and F135S variants analyzed in this study completely lost inward and outward currents when expressed alone and exhibited a clear DN effect when co-expressed with WT channels. In evaluations of intracellular trafficking, these variants showed a significant decrease in cell membrane accumulation, confirming severe intracellular trafficking defects. This suggests a typical DN mechanism in which mutant channel proteins are trapped by quality control systems in the endoplasmic reticulum (ER) and remain intracellular while forming heteromultimers with WT subunits, thereby depleting the number of functional channels on the cell membrane^12)^. Previous studies have also classified and evaluated ATS-causing variants based on the presence or absence of trafficking defects, and our present results are consistent with this framework^13)^.

### Dominant-Negative Effect with Normal Trafficking (76insT, N190I)

In contrast, the 76insT and N190I variants exerted a strong DN effect against WT, completely abolishing channel currents, even though their trafficking to the cell membrane was normally preserved. These results suggest that although the mutant subunits reach the cell membrane and form tetramers with WT subunits, they cause structural disruption of the ion conduction pathway (pore) or defective interactions with membrane lipids essential for gating, such as phosphatidylinositol 4,5-bisphosphate (PIP_2_)^19–21)^. The inclusion of even one mutant subunit in a tetramer (poisoning effect) inactivates the conductance of the entire channel, thereby exerting a strong DN effect. The mechanism by which many mutant channels with normal trafficking fail to open due to defective interactions with PIP_2_ is consistent with previous findings^3, 4, 12)^.

### A Unique Mechanism of "Haploinsufficiency" Presented by Q310R

A notable biophysical finding in this study is the intracellular dynamics of the Q310R variant. Although the Q310R variant completely lost it’s current when expressed alone, it did not show a DN effect when co-expressed with WT, instead presenting a typical haploinsufficiency phenotype (current density equivalent to that of the half-WT control). Furthermore, normal trafficking to the cell membrane was confirmed, and quantitative Co-IP assays demonstrated that the Q310R variant retains the ability to physically co-assemble with WT subunits and form multimers at an equivalent efficiency to WT channels. Normally, when a mutant subunit forms a multimer with WT and traffics to the cell membrane, a strong DN effect occurs if the pore function is disrupted because it inactivates the entire channel; however, Q310R contradicts this theory. As a hypothesis to explain this paradox, Q310R homotetramers may be nonfunctional, but heterotetramers comprising a mix of normal WT subunits and Q310R subunits may retain sufficient activity to limit DN suppression. While early studies have categorized Kir2.1 mutations into those showing a DN effect and those showing pure haploinsufficiency^11)^, these observations suggest a potential distinct mechanism of LOF in which the impact of subunit interactions on pore function is not uniform but varies depending on the mutation site.

### Partial Functional Maintenance Derived from Structural Features (I72T)

In this study, the I72T variant exhibited no trafficking defects and was the only analyzed mutation to retain partial channel function (42% inward current and 14% outward current compared to WT alone). The 72nd isoleucine (I72) in *KCNJ2* is located at the interface transitioning to the first transmembrane domain (M1), immediately preceding the slide helix (amino acid 73 and onwards), which is crucial for channel gating and PIP_2_ affinity^22)^. It is reported that mutations at D71 and T75, which flank I72, cause complete LOF and a strong DN effect^11, 14)^. Despite its important location and the severe functional consequences of neighboring variants, I72T retained substantial channel function. This might be explained, at least in part, by the physicochemical properties of amino acids. Both the WT isoleucine and the mutant threonine are β-branched amino acids, and the steric bulk of their side chains are similar. In addition, while this substitution involves a change in classification from hydrophobic to hydrophilic, it does not introduce a new charge (both are neutral). This structural and electrostatic similarity may help preserve the local architecture around the slide helix, contributing to multimer formation at the cell membrane and partial retention of pore function. These findings raise the possibility that the residual current contributes to clinical variability^15)^.

### Functional Mechanisms and Trends in Clinical Phenotypes

Comparing the functional characteristics of the identified variants with the clinical backgrounds of the respective patients suggest a possible association between functional mechanism and clinical phenotype. In the group showing a strong DN effect (103_104del, F135S, 76insT, and N190I), dysmorphism characteristic of ATS was observed in seven out of eight cases, indicating a tendency for phenotypes to be consistent. Conversely, in cases without a DN effect, such as Q310R, which showed haploinsufficiency, and I72T, which showed partial functional maintenance, dysmorphism was absent, and there was a tendency for the classical triad of ATS to be incomplete. These findings suggest that the residual function of the channel (the presence or absence of a DN effect) may influence incomplete penetrance and phenotypic variability. However, since the accumulation of clinical data in this study is limited to a small number of cases for each variant, it is difficult to draw definitive conclusions regarding this phenotypic difference. Further detailed investigations with a larger sample of cases across multiple centers using large-scale registries are awaited to determine the impact of functional severity on clinical prognosis.

### Limitations

There are several limitations to consider in interpreting this study. First, the electrophysiological and cell biological functional analyses in this study were conducted using a heterologous expression system with human embryonic kidney (HEK293) cells. HEK293 cells do not perfectly replicate the intracellular environment, such as the localization of scaffolding proteins specific to cardiomyocytes (e.g., SAP97) or specific membrane lipids (e.g., PIP_2_) essential for regulating Kir2.1 channel function^23)^. Due to these limitations of the HEK293 cell system, future studies using patient-derived induced pluripotent stem cell-derived cardiomyocytes (iPSC-CMs) are warranted, as they are becoming the standard for disease modeling studies^24)^. Second, regarding the Q310R variant, single-channel recordings were not performed to directly assess the extent to which heterotetrameric channels composed of Q310R and WT subunits retain single-channel conductance and open probability at the cell membrane. Third, this study primarily focused on elucidating fundamental molecular mechanisms, and the number of applicable cases (n) for each variant examined for genotype-phenotype correlation is limited. Therefore, the relationship between the residual function of the channel and the severity of the clinical phenotype discussed above remains a hypothesis and cannot support definitive statistical conclusions.

## Data Availability

The data underlying this article will be shared on reasonable request to the corresponding author.

## Ethics Statement

The data are available upon reasonable request. This study was approved by the institutional review committee of Shiga University of Medical Science (approval number G2011-128) and was approved by the National Cerebral and Cardiovascular Center Ethics Committee (R22006-10, M29-167-11). All subjects provided informed consent.

## Declaration of AI and AI-assisted technologies in the writing process

During the preparation of this work, the author used Gemini in order to improve the English language and readability. After using this tool/service, the author reviewed and edited the content as needed and takes full responsibility for the content of the publication.

## Acknowledgments

The authors would like to thank Arisa Ikeda, Kazu Toyooka, and Kohki Nukada for their excellent technical assistance with the genetic testing.

## Non-standard Abbreviations and Acronyms.

ATS: Andersen-Tawil syndrome
LOF: loss-of-function
DN: dominant-negative
WT: wild-type

